# Impact of antimicrobial resistance measures and the emergence of COVID-19 on antimicrobial use throughout the Japanese population: A retrospective cohort study using a national claims database

**DOI:** 10.1101/2025.06.12.25329180

**Authors:** Ryuji Koizumi, Shinya Tsuzuki, Yusuke Asai, Kensuke Aoyagi, Norio Ohmagari

## Abstract

**Objective:** This study examined the impact of Japan’s National Action Plan on Antimicrobial Resistance (NAP) and the COVID-19 pandemic on national trends in antimicrobial consumption (AMC) for major infectious diseases.

**Methods:** This retrospective cohort study analyzed claims data for the Japanese population from 2013 to 2020. AMC was expressed as defined daily doses per 1,000 inhabitants per day. The target diseases included upper respiratory infection (URI), otitis media, pneumonia, diarrhea, urinary tract infection, skin and soft tissue infection, and sexually transmitted infection. Seasonally adjusted interrupted time-series analyses were conducted to assess the impact of NAP publication in 2016 and the COVID-19 outbreak in 2020 on the numbers of medically attended cases and antimicrobial prescription rates.

**Results:** Total AMC declined from 2016 to 2019, and decreased further in 2020. There was a significant decrease in medically attended cases in 2020 when compared to the counterfactual scenario in which COVID-19 did not occur. AMC was positively correlated with the number of medically attended cases. Total antimicrobial prescription rates decreased after NAP publication and the COVID-19 outbreak. Among the diseases, prescription rates for URI, otitis media, and pneumonia significantly decreased after both events. However, the prescription rate for diarrhea decreased after NAP publication but increased after COVID-19. No significant trends were detected for other diseases.

**Conclusions:** Antimicrobial prescriptions for respiratory infections steadily decreased after NAP publication, which may indicate the effects of antimicrobial stewardship activities. Continued monitoring is needed to clarify the long-term effects of the COVID-19 pandemic on antimicrobial use.

## Introduction

Antimicrobial resistance (AMR) is an urgent global health threat [1]. Based on the framework established by the World Health Organization, many countries have developed national action plans (NAPs) to counter AMR [2]. The Japanese government published its first NAP on AMR (NAP1) in April 2016, and released its updated version (NAP2) in April 2023 [3]. These plans describe AMR indicators and measures, such as antimicrobial surveillance, educational activities, and financial incentives to encourage appropriate infection prevention and control practices. Research centers have also been established to promote and evaluate these measures [3].

Japan’s NAPs emphasize appropriate antimicrobial use, with measures that target the reduction of overuse/misuse and the promotion of diagnosis-based prescriptions [3]. Although Japan’s antimicrobial consumption (AMC) in the human health sector is not quantitatively high when compared to other countries, there is a tendency to overuse cephalosporins, macrolides, and fluoroquinolones [4]. Accordingly, reduction targets for these antimicrobials were set in both NAPs. In response, some hospitals established antimicrobial stewardship teams whose efforts have led to reductions in broad-spectrum antimicrobial use [5].

Approximately 80% of antimicrobials in Japan are used to treat respiratory infections and diarrhea [6]. A claims database study found that, despite a declining trend, antimicrobials were prescribed to approximately 30% of respiratory infection cases in 2017 [7]. Antimicrobials were also reportedly prescribed to patients with nonbacterial diarrhea [8]. Other studies have detected regional differences in AMC, and noted an association between the population-adjusted number of upper respiratory infection (URI) diagnoses and AMC [6,9]. Efforts have been made in Japan to improve antimicrobial stewardship, especially for the common cold and diarrhea [3,10]. The Japanese government has also introduced financial incentives to discourage unnecessary antimicrobial prescriptions for pediatric patients [11].

However, the abrupt onset of the COVID-19 pandemic in 2020 had profound effects on healthcare, and may have impacted AMC trends and AMR measures. The pandemic affected the treatment of other infectious diseases, with studies reporting marked reductions in AMC [3,4,12]. It has been suggested that these reductions were not due to improvements in antimicrobial use, but were instead an indirect effect of non-pharmaceutical interventions such as movement restrictions [13]. The enhancement of general infectious disease control measures to prevent COVID-19 transmission may also have led to widespread reductions in other infections. Therefore, it is uncertain if the reported reductions in AMC were due to changes in prescribing patterns or a decline in medically attended cases.

The impact of NAP1 and the COVID-19 pandemic—which we hypothesize to have substantial influence on AMC—remains unclear. This study analyzed AMC trends in the Japanese population from 2013 to 2020, and quantified the impact of these two events on the numbers of medically attended cases for major infectious diseases and their antimicrobial prescription rates.

## Methods

### Study design and patient selection

This retrospective observational cohort study was conducted using claims data for the entire population of Japan from 2013 to 2020. AMC was evaluated for all patients included in the database. We also analyzed the numbers of medically attended cases and antimicrobial prescription rates for the following target diseases: URI, otitis media (OM), pneumonia, diarrhea, urinary tract infection (UTI), skin and soft tissue infection (SSTI), and sexually transmitted infection (STI).

This study was approved by the Ethics Committee of the National Center for Global Health and Medicine Hospital (NCGM-G-003098-02).

### Data source and settings

Data were obtained from the National Database of Health Insurance Claims and Specific Health Checkups of Japan (NDB) [14]. The NDB encompasses information (including diagnoses, drug prescriptions, and medical procedures) on all patients who received insurance-covered medical treatment, regardless of insurance type. As all of Japan’s residents are covered under its universal health insurance system, the NDB can be considered to contain data for almost the entire population.

National AMC was estimated as the defined daily doses per 1,000 inhabitants per day (DID) [15]. Systemic antimicrobials administered orally or parenterally were identified using the Anatomical Therapeutic Chemical code J01 [15]. We included antimicrobial prescriptions from both inpatient and outpatient care at clinics and hospitals. In addition to total AMC, we also analyzed AMC according to the following antimicrobial categories specified as targets in Japan’s NAPs: oral cephalosporins, oral macrolides, oral fluoroquinolones, other oral antimicrobials, and parenteral antimicrobials [3]. DID was calculated based on the population size in October 1 of each year [16].

We identified medically attended cases as individuals who underwent initial consultations with a recorded diagnosis of any target disease in their insurance claims. Prescription rates were analyzed as the proportion of these patients who were prescribed antimicrobials. The target diseases were identified using recorded disease names, which were based on the corresponding International Classification of Diseases, 10th Revision codes. The selection of disease names was reviewed by several physicians and pharmacists specializing in infectious diseases.

### Statistical analysis

We analyzed the trends in the (i) number of medically attended cases and (ii) prescription rate (expressed as the number of prescriptions per medically attended case per month) for each target disease using a seasonally adjusted interrupted time-series analysis (saITSA) [17]. Seasonality was assessed as a 12-month period.

First, the saITSA for the number of medically attended cases used the following equation:

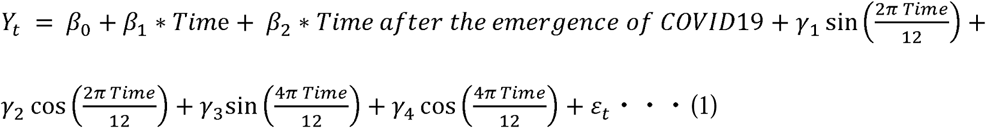

The variables and coefficients in this equation are described in **Table 1**. The data were assumed to follow a Poisson distribution, and the “tsModel” package, “Epi” package, and “glm” function were used for this saITSA.

**Table 1.** Definitions of the variables and coefficients used in the seasonally adjusted.

| **Number of medically attended cases** |  |
| --- | --- |
| Variables/Coefficients | Definition |
| $Y_t$ | Number of medically attended cases of each target disease at time $t$ (months) |
| $Time$ | Time elapsed since January 2013 (months) |
| $Time\ after\ the\ emergence\ of\ COVID-19$ | Time elapsed since January 2020, when the COVID-19 outbreak began in Japan (months) |
| $\beta_0$ | Baseline number of medically attended cases at the start of the observation period ($t=0$) |
| $\beta_1$ | Long-term temporal trend |
| $\beta_2$ | Change in trend following the emergence of the COVID-19 pandemic |
| $\gamma_1, \gamma_2$ | Coefficients representing the amplitude and phase, respectively, of the annual seasonal cycle |
| $\gamma_3, \gamma_4$ | Coefficients representing the amplitude and phase, respectively, of the biannual cycle |
| $\epsilon_t$ | Error term for unexplained variations not captured by the model |
| **Prescription rate** |  |
| Variables/Coefficients | Definition |
| $Z_t$ | Antimicrobial prescription rate of each target disease at time $t$ (months) |
| $Time$ | Time elapsed since January 2013 (months) |
| $Time\ after\ NAP1\ publication$ | Time elapsed since April 2016, when NAP1 was published in Japan (months) |
| $Time\ after\ the\ emergence\ of\ COVID-19$ | Time elapsed since January 2020, when the COVID-19 outbreak began in Japan (months) |
| $\mu_0$ | Baseline antimicrobial prescription rate at the start of the observation period ($t=0$) |
| $\mu_1$ | Long-term temporal trend |
| $\mu_2$ | Change in trend following the publication of NAP1 |
| $\mu_3$ | Change in trend following the emergence of the COVID-19 pandemic |
| $\nu_1, \nu_2$ | Coefficients representing the amplitude and phase, respectively, of the annual seasonal cycle |
| $\nu_3, \nu_4$ | Coefficients representing the amplitude and phase, respectively, of the biannual cycle |
| $\delta_t$ | Error term for unexplained variations not captured by the model |

NAP1, National Action Plan on Antimicrobial Resistance, 2016–2020.

Next, the saITSA for prescription rate used the following equation:

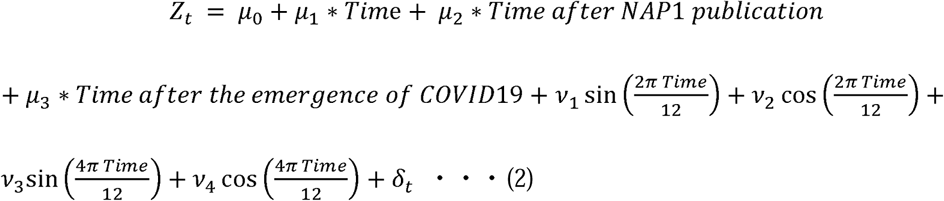

The variables and coefficients in this equation are also described in **Table 1**. The data were assumed to follow a beta distribution, and the model used a logit link function. The “betareg” function was used for this saITSA.

The 95% confidence intervals for the numbers of medically attended cases and prescription rates were estimated using the bootstrap method. We applied the “boot” package, and performed 1,000 bootstrap resamples for each simulation.

*P* values below 0.05 (two-tailed) were considered significant. All statistical analyses were conducted using R ver. 4.4.0 (R Foundation for Statistical Computing, Vienna, Austria).

## Results

### Antimicrobial consumption

The AMC of the Japanese population from 2013 to 2020 is shown in **Figure 1**. Following the publication of NAP1 in April 2016, total AMC declined from 14.5 DID in 2016 to 13.1 DID in 2019, and decreased further to 10.4 DID in 2020. When analyzed according to administration route, oral antimicrobials generally showed a declining trend. In particular, cephalosporins, macrolides, and fluoroquinolones demonstrated consistent decreases over time (**Supplementary Table 1**). In contrast, parenteral antimicrobials did not show any notable changes during the study period.

**Figure 1.**
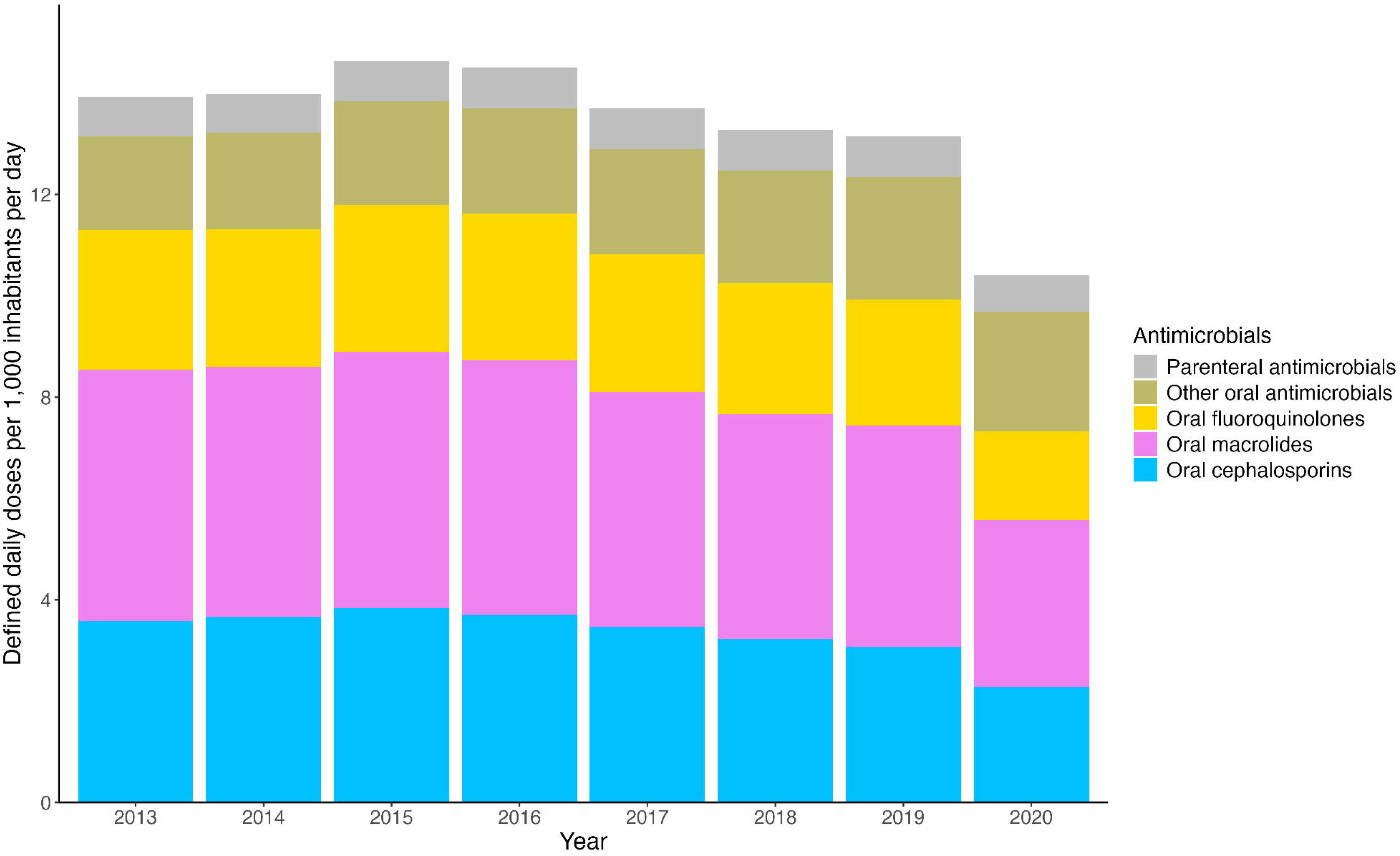
Trends in antimicrobial consumption in the Japanese population from 2013 to 2020. Antimicrobial categories correspond to the targets specified in Japan’s National Action Plan on Antimicrobial Resistance. The antimicrobials were classified using their corresponding Anatomical Therapeutic Chemical codes: cephalosporins (J01DB/C/D), macrolides (J01FA), fluoroquinolones (J01MA), other oral antimicrobials (other J01 codes; oral), and parenteral antimicrobials (other J01 codes; parenteral).

### Numbers of medically attended cases

Figure 2 presents the trends in the number of medically attended cases based on the saITSA. There were no substantial changes in the total number of medically attended cases from 2013 to 2019. However, the total and disease-specific numbers of medically attended cases demonstrated significant changes after the start of the COVID-19 pandemic in January 2020 when compared to the counterfactual scenario in which COVID-19 did not occur (**Table 2**).

**Figure 2.**
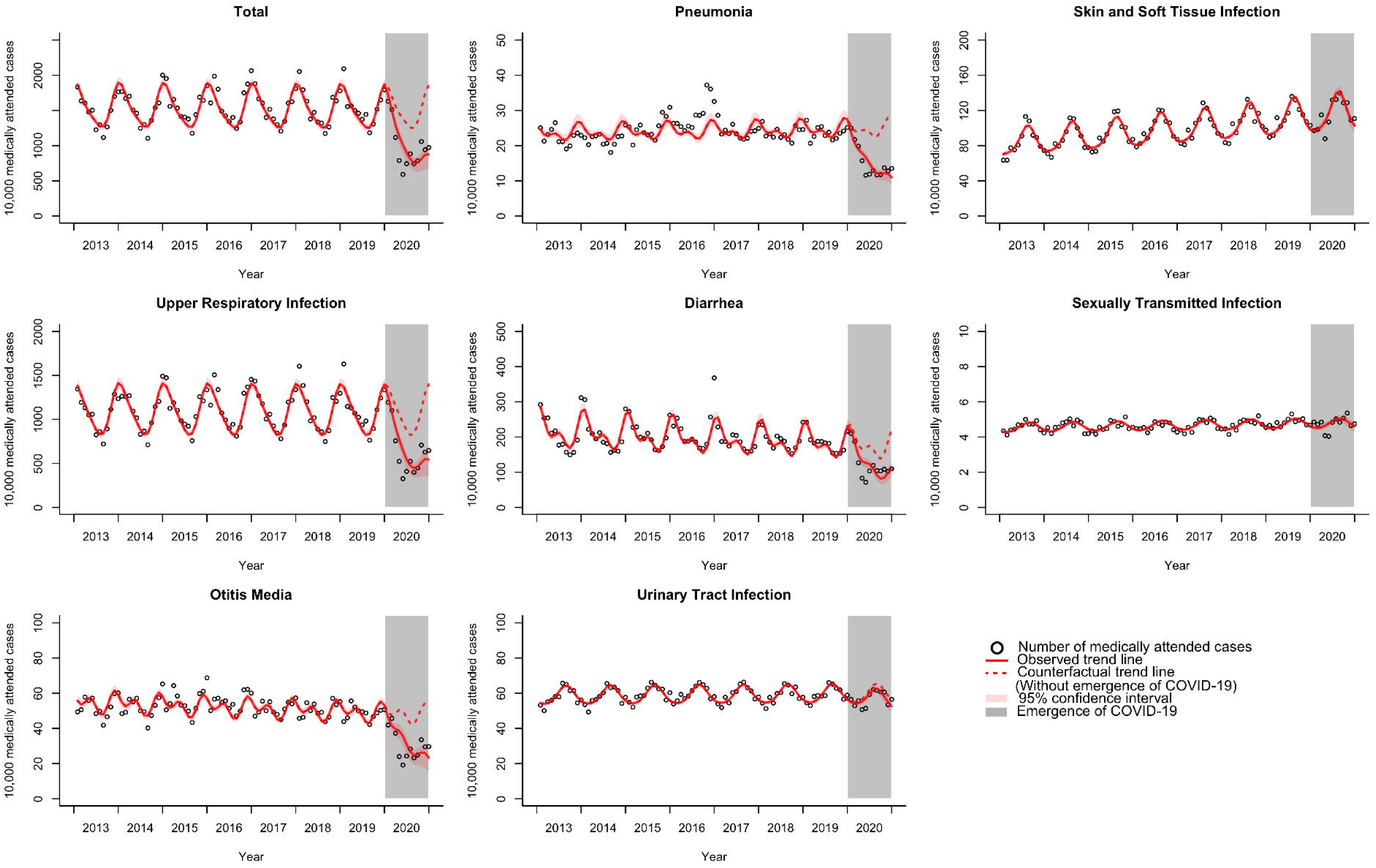
Interrupted time-series analysis of medically attended cases according to target disease.

**Table 2.**
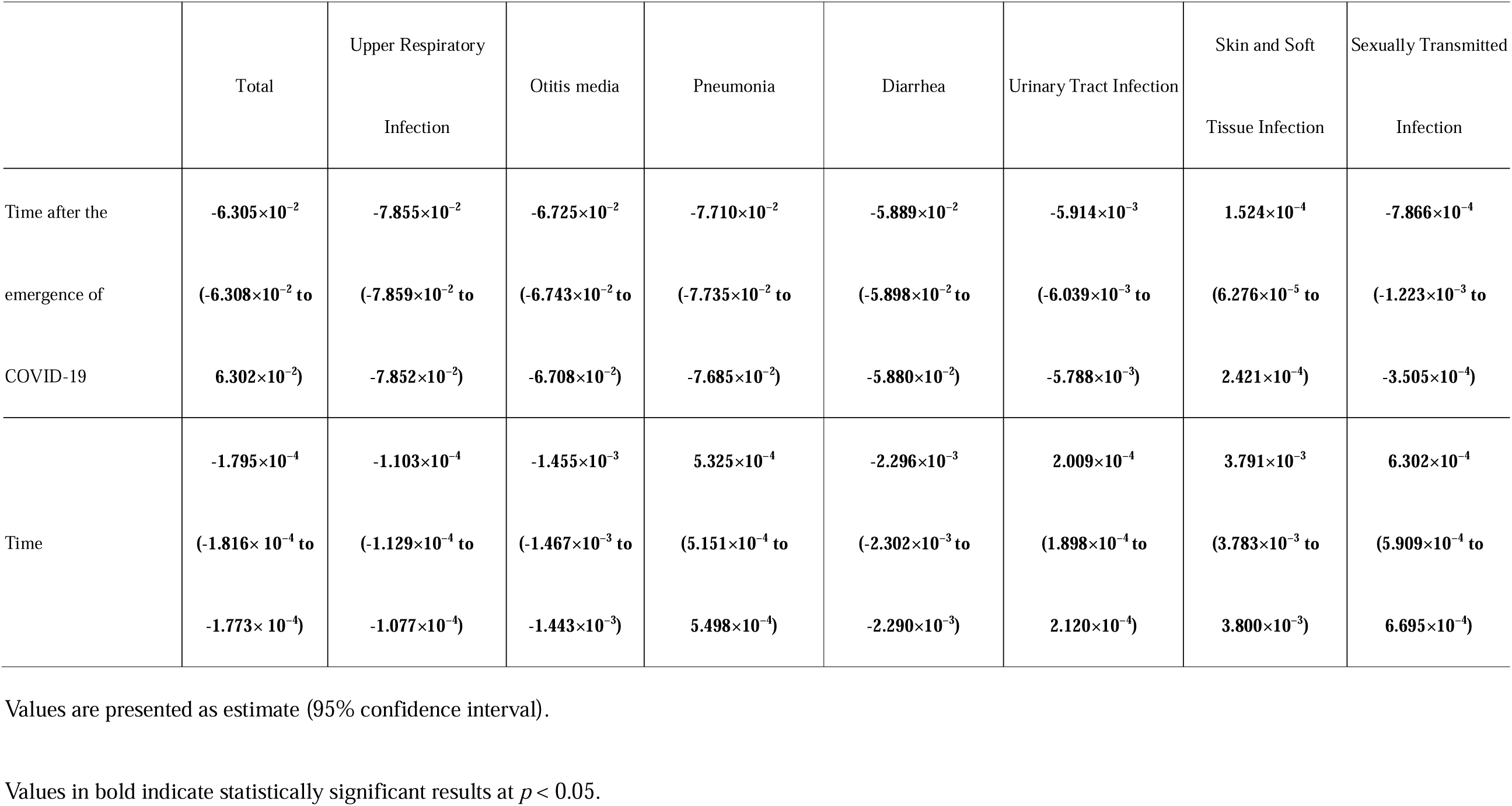
Results of the interrupted time-series analysis of medically attended cases (Poisson distribution)

|  | Total | Upper Respiratory<br>Infection | Otitis media | Pneumonia | Diarrhea | Urinary Tract Infection | Skin and Soft<br>Tissue Infection | Sexually Transmitted<br>Infection |
| --- | --- | --- | --- | --- | --- | --- | --- | --- |
| Time after the<br>emergence of<br>COVID-19 | **-6.305×10<sup>-2</sup>**<br>(-6.308×10<sup>-2</sup> to<br>**6.302×10<sup>-2</sup>**) | **-7.855×10<sup>-2</sup>**<br>(-7.859×10<sup>-2</sup> to<br>**-7.852×10<sup>-2</sup>**) | **-6.725×10<sup>-2</sup>**<br>(-6.743×10<sup>-2</sup> to<br>**-6.708×10<sup>-2</sup>**) | **-7.710×10<sup>-2</sup>**<br>(-7.735×10<sup>-2</sup> to<br>**-7.685×10<sup>-2</sup>**) | **-5.889×10<sup>-2</sup>**<br>(-5.898×10<sup>-2</sup> to<br>**-5.880×10<sup>-2</sup>**) | **-5.914×10<sup>-3</sup>**<br>(-6.039×10<sup>-3</sup> to<br>**-5.788×10<sup>-3</sup>**) | **1.524×10<sup>-4</sup>**<br>(6.276×10<sup>-5</sup> to<br>**2.421×10<sup>-4</sup>**) | **-7.866×10<sup>-4</sup>**<br>(-1.223×10<sup>-3</sup> to<br>**-3.505×10<sup>-4</sup>**) |
| Time | **-1.795×10<sup>-4</sup>**<br>(-1.816×10<sup>-4</sup> to<br>**-1.773×10<sup>-4</sup>**) | **-1.103×10<sup>-4</sup>**<br>(-1.129×10<sup>-4</sup> to<br>**-1.077×10<sup>-4</sup>**) | **-1.455×10<sup>-3</sup>**<br>(-1.467×10<sup>-3</sup> to<br>**-1.443×10<sup>-3</sup>**) | **5.325×10<sup>-4</sup>**<br>(5.151×10<sup>-4</sup> to<br>**5.498×10<sup>-4</sup>**) | **-2.296×10<sup>-3</sup>**<br>(-2.302×10<sup>-3</sup> to<br>**-2.290×10<sup>-3</sup>**) | **2.009×10<sup>-4</sup>**<br>(1.898×10<sup>-4</sup> to<br>**2.120×10<sup>-4</sup>**) | **3.791×10<sup>-3</sup>**<br>(3.783×10<sup>-3</sup> to<br>**3.800×10<sup>-3</sup>**) | **6.302×10<sup>-4</sup>**<br>(5.909×10<sup>-4</sup> to<br>**6.695×10<sup>-4</sup>**) |

Values are presented as estimate (95% confidence interval).
Values in bold indicate statistically significant results at $p < 0.05$ .

Among the target diseases, SSTI, STI, and UTI did not decline in 2020, with median change rates (calculated by comparing the annual median values from 2013–2019 to 2020) of +16.5%, +3.7%, and +3.2%, respectively. In contrast, we observed reductions in medically attended cases for URI (median change rate: -48.0%), diarrhea (-43.4%), OM (-43.5%), and pneumonia (-43.8%). URI and diarrhea were the most common medically attended cases in our study population. The median number of URI cases was 11,068,993.5 in 2013–2019, but decreased to 5,759,782 in 2020. Similarly, the median number of diarrhea cases was 1,880,289 in 2013–2019, but decreased to 1,065,005.5 in 2020. The total monthly number of medically attended cases was correlated with total monthly DID, and both variables underwent a decrease in 2020 (**Supplementary Figure 1**).

### Antimicrobial prescription rates

Figure 3 presents the trends in antimicrobial prescription rates based on the saITSA. The total prescription rate showed a decreasing trend in each year after the publication of NAP1 in April 2016. When analyzed according to target disease, the prescription rates for URI, OM, and pneumonia had gradually decreased after NAP1 publication, but underwent a more pronounced decline following the emergence of COVID-19. The prescription rate for URI had the largest decrease from 57.8% in 2013 to 38.2% in 2020. While the prescription rate for diarrhea decreased after NAP1 publication, it began to increase slightly after the emergence of COVID-19. The prescription rates for the other target diseases showed no significant changes in 2020 (**Table 3**).

**Figure 3.**
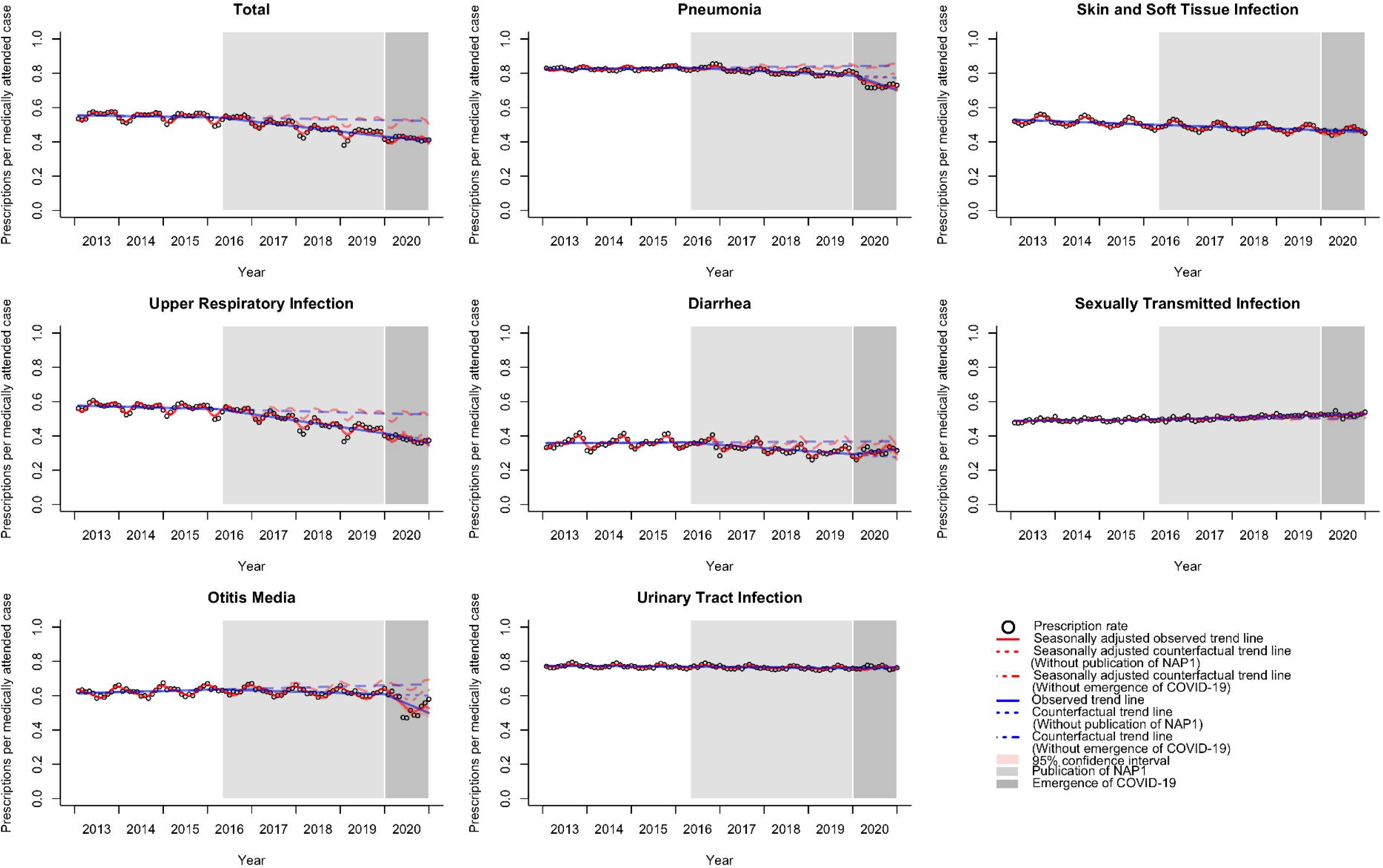
Interrupted time-series analysis of antimicrobial prescription rates according to target disease. NAP1, National Action Plan on Antimicrobial Resistance, 2016–2020.

**Table 3.** Results of the interrupted time-series analysis of antimicrobial prescription rates (beta distribution)

|  | Total | Upper Respiratory<br>Infection | Otitis media | Pneumonia | Diarrhea | Urinary Tract Infection | Skin and Soft<br>Tissue Infection | Sexually Transmitted<br>Infection |
| --- | --- | --- | --- | --- | --- | --- | --- | --- |
| Time after NAP1<br>publication | **-7.783×10<sup>-3</sup>**<br>(-9.315×10<sup>-3</sup> to<br>-6.250×10<sup>-3</sup>) | **-9.637×10<sup>-3</sup>**<br>(-1.161×10<sup>-2</sup> to<br>-7.664×10<sup>-3</sup>) | **-4.804×10<sup>-3</sup>**<br>(-7.038×10<sup>-3</sup> to<br>-2.569×10<sup>-3</sup>) | **-7.336×10<sup>-3</sup>**<br>(-9.220×10<sup>-3</sup> to<br>-5.451×10<sup>-3</sup>) | **-6.601×10<sup>-3</sup>**<br>(-8.810×10<sup>-3</sup> to<br>-4.391×10<sup>-3</sup>) | -3.666×10<sup>-4</sup><br>(-1.534×10<sup>-3</sup> to<br>8.006×10<sup>-4</sup>) | **1.186×10<sup>-3</sup>**<br>(3.531×10<sup>-4</sup> to<br>2.019×10<sup>-3</sup>) | **1.959×10<sup>-3</sup>**<br>(8.460×10<sup>-4</sup> to<br>3.074×10<sup>-3</sup>) |
| Time after the<br>emergence of<br>COVID-19 | -1.275×10<sup>-3</sup><br>(-6.071×10<sup>-3</sup> to<br>3.521×10<sup>-3</sup>) | **-1.229×10<sup>-2</sup>** -<br>(-1.853×10<sup>-2</sup> to<br>-6.050×10<sup>-3</sup>) | **-3.583×10<sup>-2</sup>**<br>(-4.265×10<sup>-2</sup> to<br>-2.900×10<sup>-2</sup>) | **-3.585×10<sup>-2</sup>**<br>(-4.119×10<sup>-2</sup> to<br>-3.052×10<sup>-2</sup>) | **1.336×10<sup>-2</sup>**<br>(6.351×10<sup>-3</sup> to<br>2.036×10<sup>-2</sup>) | 3.323×10<sup>-3</sup><br>(-2.863×10<sup>-4</sup> to<br>6.933×10<sup>-3</sup>) | -2.194×10<sup>-3</sup><br>(-4.787×10<sup>-3</sup> to<br>3.989×10<sup>-4</sup>) | -2.443×10<sup>-3</sup><br>(-5.910×10<sup>-3</sup> to<br>1.025×10<sup>-3</sup>) |
| Time | **-1.920×10<sup>-3</sup>**<br>(-2.886×10<sup>-3</sup> to | **-2.777×10<sup>-3</sup>**<br>(-4.022×10<sup>-3</sup> to | **2.261×10<sup>-3</sup>**<br>(8.517×10<sup>-4</sup> | 1.115 ×10<sup>-3</sup><br>(-8.505×10<sup>-5</sup> to | -6.681×10<sup>-5</sup><br>(-1.451×10<sup>-3</sup> to | **-8.627×10<sup>-4</sup>**<br>(-1.602×10<sup>-3</sup> to | **-3.623×10<sup>-3</sup>**<br>(-4.148×10<sup>-3</sup> to | 5.230×10<sup>-4</sup><br>(-1.796×10<sup>-4</sup> to |
|  | **-9.532×10<sup>-4</sup>**) | **-1.532×10<sup>-3</sup>**) | **3.670×10<sup>-3</sup>**) | 2.315×10<sup>-3</sup>) | 1.317×10<sup>-3</sup>) | **-1.238×10<sup>-4</sup>**) | **-3.097×10<sup>-3</sup>**) | 1.225×10<sup>-3</sup>) |

Values are presented as estimate (95% confidence interval).
Values in bold indicate statistically significant results at $p < 0.05$ .
NAP1, National Action Plan on Antimicrobial Resistance, 2016–2020.

## Discussion

To our knowledge, this is the first study to comprehensively analyze the temporal trends in antimicrobial use for major infectious diseases across the Japanese population. In contrast to previous research that primarily focused on hospitals or specific regions, our study analyzed AMC in outpatients and inpatients treated at all clinics and hospitals throughout the country [18,19]. The publication of NAP1 and the emergence of COVID-19 likely influenced infectious disease management in Japan, and our findings shed light on the potential impact of these two events on healthcare-seeking behavior and antimicrobial prescribing practices.

Our study observed a correlation between the monthly number of medically attended cases and AMC, both of which declined in 2020. We posit that the reduction in medically attended cases influenced the decrease in AMC. There were no substantial changes in the number of medically attended cases for the target diseases until 2019. While the numbers of diarrhea cases and SSTI cases had been on gradual downward and upward trends, respectively, no outbreaks were reported between 2013 and 2019. These trends may have been due to shifts in diagnostic patterns influenced by the implementation of antimicrobial stewardship initiatives and the dissemination of clinical guidelines that promoted more careful evaluations before diagnoses [3,10,20]. Next, our study observed sudden reductions in medically attended pneumonia, OM, and URI cases in 2020. An analysis of national infectious disease surveillance data found that the number of consultations for respiratory-transmitted bacterial infections in Japan significantly decreased in 2020, which may have been driven by non-pharmaceutical interventions for COVID-19 [21]. Similar phenomena have also been observed in other countries [13,22]. The strengthening of infection control measures and decline in individuals seeking medical care for minor illnesses could have influenced our observed trends [23,24]. Our study also found that the number of medically attended SSTI cases continued to increase in 2020, which may have been influenced by the dissemination of guidelines on skin diseases and infections. In contrast, the number of medically attended UTI cases showed no notable changes throughout the study period. UTIs are generally not epidemic in nature, and their incidence rates are unlikely to be influenced by NAP1. Although studies in Japan and the US have reported a rise in syphilis cases after the lifting of COVID-19 restrictions, our analysis found no notable changes in STI cases [25,26].

Next, we found that antimicrobial prescription rates significantly decreased for URI, OM, pneumonia, and diarrhea from the publication of NAP1 in April 2016 until 2019. These observations were likely due to the fact that Japan’s AMR countermeasures mainly focus on respiratory infections and diarrhea [3,10,11]. Specifically, initiatives targeting these diseases (e.g., clinical guidelines, financial incentives, and educational programs) may have collectively contributed to the observed reductions. Previous studies have reported antimicrobial prescription rates of approximately 30% for both URI and diarrhea, which were lower than our observed rates [7,8]. This discrepancy may stem from the limited data sources used in previous research. Our ecological study used nationwide data, which offered a more comprehensive perspective; however, this approach precluded the consideration of individual-level factors.

After the emergence of COVID-19 in 2020, antimicrobial prescriptions significantly decreased for URI, OM, and pneumonia; significantly increased for diarrhea; and showed no changes for UTI, SSTI, and STI. However, our results only reflect the initial phase of the pandemic, and definitive conclusions cannot be drawn. One possible explanation for the reduced prescriptions for URI, OM, and pneumonia could be that the widespread use of COVID-19 testing allowed for more precise identification of these diseases [27,28]. Furthermore, the overall decrease in medically attended cases would have allowed physicians to allocate more time for each patient, thereby facilitating more accurate diagnoses. On the other hand, the relatively high proportion of non-contact consultations for common diseases such as diarrhea may have made diagnoses less certain, which could have contributed to an increase in antimicrobial prescriptions. Reports from the US have also suggested that the COVID-19 pandemic led to delays in AMR testing and hindered the diagnosis of other infections, potentially driving inappropriate antimicrobial use [29]. In addition, a high incidence of pathogenic *Escherichia coli*-associated gastroenteritis was reported among pediatric patients in Japan in 2020, which may also have influenced this trend [30]. While we did not observe any changes in antimicrobial prescriptions for UTIs, SSTIs, and STIs, it is possible that the outbreak did not greatly affect the incidence of these diseases during the study period. It is also possible that a reluctance to seek care for minor symptoms led to an increase in the proportion of relatively severe cases. The influence of the COVID-19 pandemic on antimicrobial prescription rates has yet to be fully elucidated, and there is a need for continued long-term monitoring and evaluations.

This study has several limitations. First, the analysis was conducted using insurance claims data, which did not include information on self-medication. However, the majority of essential healthcare services are covered by health insurance, which has universal enrollment in Japan. Therefore, this limitation is not expected to have a substantial impact on our results. Second, our analytical models may contain residual and unmeasured confounding due to the lack of other data sources, including potentially influential events (e.g., clinical guidelines) that occurred after April 2016. However, we assumed that relevant events would generally represent activities based on NAP1 or COVID-19, and as such would be encompassed in our analytical models. Finally, the impact of COVID-19 was only assessed for one year due to the lack of available data, which prevented any insight into its long-term effects on antimicrobial use.

In conclusion, our study indicates that the introduction of NAP1 contributed to a decrease in the overall antimicrobial prescription rate that was independent of the COVID-19 outbreak.

However, the pandemic significantly reduced the number of medically attended cases for various diseases, which prompted a further decrease in AMC in 2020. Continued monitoring is needed to clarify the long-term effects of the pandemic and NAP2 on antimicrobial use in Japan.

## Data Availability

Data from this study cannot be shared due to legal and administrative restrictions. Data may be directly acquired from the Ministry of Health, Labour and Welfare of Japan for researchers who meet the criteria for access to confidential data. Please contact the corresponding author for further information.

## Contributors

RK, ST, and YA conceived and designed the study. RK and KA curated and analyzed the data. ST and YA validated the results. RK, ST, and YA interpreted the results. RK wrote the first draft of the manuscript, and all other authors critically reviewed and revised the manuscript for important intellectual content. NO supervised the study. All authors have read and agreed to the final version of the manuscript.

## Acknowledgments

We would like to express our sincere gratitude to Dr. Fujitomo and Dr. Hikida from the National Center for Global Health and Medicine for their invaluable advice on this study.

## References

[1] Global burden of bacterial antimicrobial resistance 1990-2021: a systematic analysis with forecasts to 2050. Lancet 2024;404:1199–226. 10.1016/s0140-6736(24)01867-1.

[2] World Health Organization. Global action plan on antimicrobial resistance 2016. https://www.who.int/publications/i/item/9789241509763 (accessed June 12, 2025).

[3] World Health Organization. Japan’s AMR response 2013-2025: developing, implementing and evaluating national AMR action plans n.d. https://www.who.int/publications/i/item/9789290620792 (accessed June 12, 2025).

[4] AMR Clinical Reference Center. Surveillance based on data from the NDB n.d. https://amrcrc.jihs.go.jp/surveillance/010/20200813164848.html (accessed June 12, 2025).

[5] Nakano Y, Hirai T, Murata M, Yasukochi H, Ura K, Sueyasu Y, et al. Impact of pharmacist-driven antimicrobial stewardship interventions in a secondary care facility in Japan: A pragmatic quasi-experimental study. J Infect Chemother 2025;31:102503. 10.1016/j.jiac.2024.08.018.

[6] Hashimoto H, Saito M, Sato J, Goda K, Mitsutake N, Kitsuregawa M, et al. Indications and classes of outpatient antibiotic prescriptions in Japan: A descriptive study using the national database of electronic health insurance claims, 2012-2015. Int J Infect Dis 2020;91:1–8. 10.1016/j.ijid.2019.11.009.

[7] Kimura Y, Fukuda H, Hayakawa K, Ide S, Ota M, Saito S, et al. Longitudinal trends of and factors associated with inappropriate antibiotic prescribing for non-bacterial acute respiratory tract infection in Japan: A retrospective claims database study, 2012-2017. PLoS One 2019;14:e0223835. 10.1371/journal.pone.0223835.

[8] Ono A, Aoyagi K, Muraki Y, Asai Y, Tsuzuki S, Koizumi R, et al. Trends in healthcare visits and antimicrobial prescriptions for acute infectious diarrhea in individuals aged 65 years or younger in Japan from 2013 to 2018 based on administrative claims database: a retrospective observational study. BMC Infect Dis 2021;21:983. 10.1186/s12879-021-06688-2.

[9] Kitano T, Tsuzuki S, Koizumi R, Aoyagi K, Asai Y, Kusama Y, et al. Factors Associated with Geographical Variability of Antimicrobial Use in Japan. Infect Dis Ther 2023;12:2745–55. 10.1007/s40121-023-00893-z.

[10] Ministry of Health, Labour and Welfare. Manual of Antimicrobial Stewardship, 3rd Edition n.d.

[11] Okubo Y, Uda K, Miyairi I. Long-term Effectiveness of Financial Incentives for Not Prescribing Unnecessary Antibiotics to Children with Acute Respiratory and Gastrointestinal Infections: A Japan’s Nationwide Quasi-Experimental Study. Clin Infect Dis 2024. 10.1093/cid/ciae577.

[12] Högberg LD, Vlahović-Palčevski V, Pereira C, Weist K, Monnet DL. Decrease in community antibiotic consumption during the COVID-19 pandemic, EU/EEA, 2020. Euro Surveill 2021;26. 10.2807/1560-7917.Es.2021.26.46.2101020.

[13] Tsuzuki S, Koizumi R, Asai Y, Ohmagari N. Trends in antimicrobial consumption: long-term impact of the COVID-19 pandemic. Clin Microbiol Infect 2024. 10.1016/j.cmi.2024.12.005.

[14] Suto M, Iba A, Sugiyama T, Kodama T, Takegami M, Taguchi R, et al. Literature Review of Studies Using the National Database of the Health Insurance Claims of Japan (NDB): Limitations and Strategies in Using the NDB for Research. Jma j 2024;7:10–20. 10.31662/jmaj.2023-0078.

[15] World Health Organization. GLASS methodology for surveillance of national antimicrobial consumption n.d. https://www.who.int/publications/i/item/9789240012639 (accessed June 12, 2025).

[16] Statistics Bureau of Japan. Statistics Bureau Home Page n.d. https://www.stat.go.jp/english/index.html (accessed June 12, 2025).

[17] Bernal JL, Cummins S, Gasparrini A. Interrupted time series regression for the evaluation of public health interventions: a tutorial. Int J Epidemiol 2017;46:348–55. 10.1093/ije/dyw098.

[18] Doyle D, Dalton B, Zhang Z, Sabuda D, Rajakumar I, Rennert-May E, et al. A quasi-experimental analysis comparing antimicrobial usage on COVID-19 and non-COVID-19 wards. Antimicrob Steward Healthc Epidemiol 2024;4:e192. 10.1017/ash.2024.417.

[19] Kim T, Moehring RW, Turner NA, Ashley ED, Crane L, Padgette P, et al. Long-term trends in the incidence of hospital-acquired carbapenem-resistant Enterobacterales and antimicrobial utilization in a network of community hospitals in the Southeastern United States from 2013 to 2023. Infect Control Hosp Epidemiol 2024;46:1–7. 10.1017/ice.2024.173.

[20] The Japanse Association for Infectious Disease,. JAID/JSC Infectious Disease Treatment Guide 2023 n.d. https://www.kansensho.or.jp/modules/journal/index.php?content_id=16 (accessed June 12, 2025).

[21] Kajihara T, Yahara K, Kamigaki T, Hirabayashi A, Hosaka Y, Kitamura N, et al. Effects of coronavirus disease 2019 on the spread of respiratory-transmitted human-to-human bacteria. J Infect 2024;89:106201. 10.1016/j.jinf.2024.106201.

[22] Tomczyk S, Taylor A, Brown A, de Kraker MEA, El-Saed A, Alshamrani M, et al. Impact of the COVID-19 pandemic on the surveillance, prevention and control of antimicrobial resistance: a global survey. J Antimicrob Chemother 2021;76:3045–58. 10.1093/jac/dkab300.

[23] Sahakyan S, Muradyan D, Giloyan A, Harutyunyan T. Factors associated with delay or avoidance of medical care during the COVID-19 pandemic in Armenia: results from a nationwide survey. BMC Health Serv Res 2024;24:20. 10.1186/s12913-023-10483-x.

[24] Horita N, Kato H, Watanabe K, Hara Y, Kobayashi N, Kaneko T. Decline in mortality due to respiratory diseases in Japan during the coronavirus disease 2019 pandemic. Respirology 2022;27:175–6. 10.1111/resp.14186.

[25] Komori A, Mori H, Xie W, Valenti S, Naito T. Rapid resurgence of syphilis in Japan after the COVID-19 pandemic: A descriptive study. PLoS One 2024;19:e0298288. 10.1371/journal.pone.0298288.

[26] Do D, Rodriguez PJ, Gratzl S, Cartwright BMG, Baker C, Stucky NL. Trends in Incidence of Syphilis Among US Adults from January 2017 to October 2024. Am J Prev Med 2025. 10.1016/j.amepre.2025.03.008.

[27] Staadegaard L, Del Riccio M, Wiegersma S, El Guerche-Séblain C, Dueger E, Akçay M, et al. The impact of the SARS-CoV-2 pandemic on global influenza surveillance: Insights from 18 National Influenza Centers based on a survey conducted between November 2021 and March 2022. Influenza Other Respir Viruses 2023;17:e13140. 10.1111/irv.13140.

[28] Arima Y, Kanou K, Arashiro T, Y KK, Otani K, Tsuchihashi Y, et al. Epidemiology of Coronavirus Disease 2019 in Japan: Descriptive Findings and Lessons Learned through Surveillance during the First Three Waves. Jma j 2021;4:198–206. 10.31662/jmaj.2021-0043.

[29] Roger PM, Challut N, Hennet MA, Lemasson A, Lesselingue D. New medical staff in the post-COVID-19 period entailed altered quality of antibiotic therapy. Infect Dis Now 2024;54:104957. 10.1016/j.idnow.2024.104957.

[30] Kashima K, Sato M, Osaka Y, Sakakida N, Kando S, Ohtsuka K, et al. An outbreak of food poisoning due to Escherichia coli serotype O7:H4 carrying astA for enteroaggregative E. coli heat-stable enterotoxin1 (EAST1). Epidemiol Infect 2021;149:e244. 10.1017/s0950268821002338.

