## Supplementary Table and Figure for "Impact of antimicrobial resistance measures and the emergence of COVID-19 on antimicrobial use throughout the Japanese population: A retrospective cohort study using a national claims database"

**Supplementary Figure 1. Monthly data plots of medically attended cases and total DID**

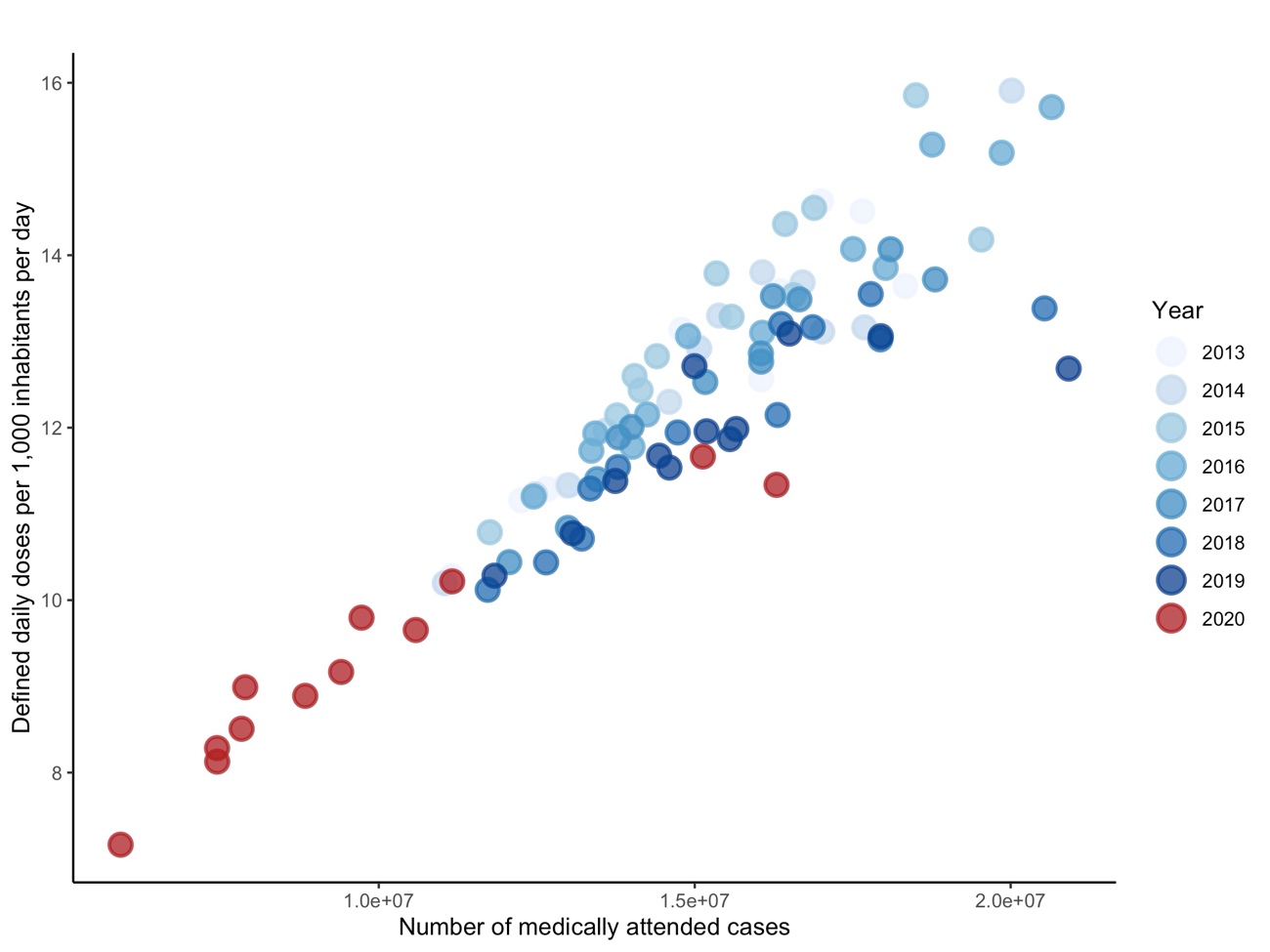

Correlation coefficient: 0.92

DID, defined daily doses per 1,000 inhabitants per day.

**Supplementary Table 1. Antimicrobial consumption in the Japanese population from 2013 to 2020**

| **Antimicrobial** | **2013** | **2014** | **2015** | **2016** | **2017** | **2018** | **2019** | **2020** |
| --- | --- | --- | --- | --- | --- | --- | --- | --- |
| Oral cephalosporins | 3.576 | 3.666 | 3.830 | 3.702 | 3.465 | 3.233 | 3.068 | 2.275 |
| Oral macrolides | 4.967 | 4.932 | 5.063 | 5.027 | 4.635 | 4.435 | 4.369 | 3.297 |
| Oral fluoroquinolones | 2.752 | 2.711 | 2.905 | 2.898 | 2.712 | 2.584 | 2.485 | 1.761 |
| Other oral antimicrobials | 1.854 | 1.907 | 2.039 | 2.081 | 2.087 | 2.219 | 2.419 | 2.359 |
| Parenteral antimicrobials | 0.779 | 0.771 | 0.794 | 0.801 | 0.804 | 0.807 | 0.805 | 0.715 |

Values are presented as the defined daily doses per 1,000 inhabitants per day.
